# ENO1 as a biomarker of breast cancer progression and metastasis – a bioinformatic approach

**DOI:** 10.1101/2023.11.01.23297919

**Authors:** Athina Giannoudis, Alistair Heath, Vijay Sharma

## Abstract

**Background:** Metabolic reprogramming is one of the hallmarks of cancer cells and many key metabolic enzymes are dysregulated. In breast cancer (BC), the commonest malignancy of women, several metabolic enzymes are overexpressed and/or overactivated. One of these is Enolase 1 (ENO1) an enzyme that catalyses glycolysis but is also involved in the regulation of multiple signalling pathways. ENO1 overexpression in BC has been linked to worse tumour prognosis and metastasis, rendering it a promising biomarker of disease progression and a potential therapeutic target.

**Methods:** Utilising available online platforms such as the KM-plotter, the ROC-plotter, the cBioPortal, the G-2-O, the MethSurvand, we performed a bioinformatic analysis to establish the prognostic and predictive effects related to ENO1 expression in breast cancer. A Network analysis was also performed using the Oncomine platform and signalling and epigenetic pathways including immune regulation constituting the hallmarks of cancer were explored. The relationship between ENO1 and the immune response was also obtained from the TISIDB portal and Spearman’s rho (r) was used to determine their correlation.

**Results:** ENO1 is overexpressed in all the analysed Oncomine, epigenetic and immune pathways in triple-negative, but not in hormone receptor-positive BCs. In HER2-positive BCs, ENO1 expression showed a mixed profile. Similarly, analysis on disease progression and histological types showed ENO1 overexpression in ductal *in situ* and invasive carcinoma, high grade tumours followed by advanced and/or metastasis and was linked to worse survival (death by 5 years). High ENO1 expression was also associated with relapse-free (RFS), distant metastasis-free (DMFS) and overall survival (OS) as analysed by the KM-plot software, irrespectively of treatment and was also related to basal subtype and to a lesser extend to HER2 and luminal B subtypes. ENO1 was underexpressed in the less invasive and with better prognosis subtypes.

**Conclusions:** Overexpression of ENO1 largely confers a worse prognosis in breast cancer and recruits a range of signalling pathways during disease progression. ENO1 expression can be utilised as a biomarker of disease progression and as a potential therapeutic target, particularly in triple-negative and invasive breast carcinomas (NST).

## Background

Breast cancer is the most prevalent malignancy in women and despite the advancements in its diagnosis and management, it is still is one of the leading causes of cancer-related death in women [1,2]. Its incidence and mortality have been reported to be 46.8% and 13.6% respectively [2,3]. Breast cancer is a very heterogeneous disease that can be classified by its histological subtype, its receptor status or its molecular phenotype [4,5]. The mainstay of oncological management of breast cancer includes endocrine therapy, human epidermal growth factor receptor 2 (HER2) targeted therapy and chemotherapy [6]. However, in addition to its heterogeneity, the presence of additional cell types such as stromal and immune cells within the tumour microenvironment makes more difficult the management of the disease [7]. There is therefore a need to identify further targets, that can be used as biomarkers of resistance and disease progression and be potentially used as therapeutic options.

Alterations in energy metabolism by cancer cells can promote tumourigenesis and it is well-established that metabolic reprogramming is one of the hallmarks of cancer cells [8,9]. There is a growing interest in exploring metabolic pathways for biomarkers and novel therapeutic targets. Glucose metabolism in cancer has received a lot of attention, mainly through the expression of glucose transporters (GLUT1/3), pyruvate dehydrogenase kinase (PDK), lactate dehydrogenase (LDH), hexokinases (HK1/2), phosphofructokinase (PFK) and its regulation by oncogenes, tumour-suppressors and transcription factors [10]. In addition, various signalling pathways, such as Notch, phosphoinositide-3-kinase (PI3K), PTEN, mammalian target of rapamycin (mTOR), and mitogen-activated protein kinase (MAPK) also interact with metabolic reprogramming [8–10]. During the last decade, despite the emerging of transporter and/or metabolic enzyme inhibitors, the efficiency of targeting glucose metabolism has proved challenging and there is a clinical need to identify and explore more promising targets. One of these targets is Enolase 1 (ENO1), a glycolytic enzyme that primarily catalyses the conversion of 2-phosphoglyceric acid to phosphor-enol-pyruvic acid during glycolysis [11,12] It is a multifunctional protein, ubiquitously expressed in most human tissues under normal and pathophysiological conditions and is found overexpressed in many cancers [11–13].

ENO1 mRNA and protein overexpression has been linked to disease progression and worse clinical outcome in lung, breast, pancreas, glioma, head and neck and colorectal cancers [13–19]. In several cancers, such as gastric, pancreatic, prostate and breast, in addition to worse outcome it has been associated to treatment resistance and in particular, chemoresistance [20–22]. In breast cancer, it’s been shown *in vitro* that silencing ENO1 inhibits the proliferation, migration and invasion of breast cancer cells [24] and in a xenograft mouse model, inhibition of ENO1 expression increased tolerance to hypoxia in tumour cells, showing also slow reduced tumour size, cell growth and increased apoptosis [25]. A recent, single-cell transcriptomic profiling of breast cancer patients, identified higher ENO1 expression in the aggressive basal subtype, compared to hormone-and/or HER2-positive subtypes and this overexpression was linked to worse relapse-free survival [26]. They also showed that depletion of ENO1 in triple-negative breast cancer cell lines halted cell proliferation, colony formation and tumour growths (3D-organoids) and increased cell death suggesting that ENO1 could be used as a therapeutic target in this aggressive subtype [26]. However, a more comprehensive data analysis will allow us to better characterise the role of ENO1 in breast cancer progression and its potential use as a targeted therapeutic agent.

The aim of this analysis is to evaluate the predictive and/or prognostic value of ENO1 as a biomarker of breast cancer progression and as a therapeutic target using a range of online bioinformatics tools. Network analysis on the breast cancer patient cohorts available on the Oncomine platform [27] will allow us to comprehensively characterise how ENO1 clusters globally with genes involved in all known cancer hallmarks, epigenetic and immune pathways.

## Methods

### Kaplan-Meier (KM) and ROC-plotter analysis

The predictive and prognostic effect of ENO1 expression at mRNA level was assessed using the Kaplan-Meier (KM) plotter (www.kmplot.com) tool and was stratified by treatment and by molecular subtype [28]. Briefly, the expression of ENO1 was divided into high and low groups by splitting the mRNA expression level at the median values. Kaplan-Meier survival analysis was performed to assess the effect on progression-free (PFS), distant metastasis-free (DMFS) and overall survival (OS). RFS is defined as the time from initial diagnosis to first recurrence of the disease. It is commonly used in trials as a surrogate marker of OS as it requires less follow up to get this measure and the information is available more quickly. DMFS is defined as the time from initial diagnosis to distant site (distant lymph nodes, lung, liver, brain) and OS is the time from initial diagnosis to death from any cause. For all the survival analysis, a log-rank p value <0.005 was considered significant.

The effect of ENO1 on treatment response was assessed through the use of the receiver operating characteristic curve (ROC) and calculating the area under the curve (AUC) using the ROC-plotter tool [29]. ROC analysis was performed for complete pathological response and 5-year RFS as short and long-term outcomes.

### Genomic alterations analysis

Alterations in the ENO1 genome were assessed using the cBioPortal tool [30,31] and the prognostic effect of these alterations was assessed by Kaplan-Meier survival analysis using the KM-plotter and the Genotype-2-Outcome (G-2-O) tools. [32]

### Oncomine Network analysis

Network analysis was performed using the Oncomine platform as previously described utilising the built-in molecular concepts [27,33]. The signalling pathways for the cancer hallmarks, including immune regulation, were based on the NanoString concepts [34] and the epigenetic signalling pathways were obtained from the EpiFactor website [35]. The assessment of the prognostic effect of methylation of the ENO1 gene was assessed using the MethSurv tool [36] by Kaplan-Meier analysis.

Briefly, ENO1 was investigated across the molecular concepts to identify clustering with different signalling pathways. Clustering of the signalling pathways with ENO1 was taken as significant at a p<0.01 and any odds ratio (OR) (Supplementary file 1). The tool specified whether the clustering occurred in the context of over-or under-expression and specified the patient subgroup in which the clustering occurred. Subgroups identified in the platform included (i) subgroups related to stage, recurrence, survival outcome, and (ii) subgroups related to the histological subtype and receptor status of the tumour.

### ENO1 and tumour-immune system interactions

Spearman correlation (r) was used to determine the relationship between ENO1 and the immune response in breast cancer. The data was obtained using the TISIDB portal [37] for all available lymphocytes, chemokines and immunomodulators.

### Ethics

The data provided on these online Bioinformatics tools is fully anonymised and ethical permission is covered by the original studies which generated the data and by the providers of the online tools. Further details are provided in the original publications of these tools [28–37]. Additional specific ethical approval was not required to conduct this study.

## Results

### ENO1 expression at mRNA level and correlation to survival

ENO1 is overexpressed at mRNA level in breast cancer in comparison to normal tissue (p=1.46×10^-16^; Figure 1A). Overexpression of ENO1 is a poor predictive marker as identified by the KM-plotter tool (Figure 1B-D) using the Affymetrix ID: 201231_s_at (ENO1, ENO1L1, MBP-1). High expression of ENO1 in breast cancer patients correlates with a decreased RFS, DMFS and OS (p<0.001 for all comparisons; Figure 1B-D respectively). Stratifying the patients by treatment status including all types of treatment showed that high ENO1 expression predicted a poor survival outcome in all patients irrespectively of treatment (Supplementary Figure 1A-C). Patient stratification by molecular subtype showed a decreased RFS, DMFS and OS in HER2-positive and basal breast cancer subtypes (p<0.05 for all comparisons; Figure 2A-C) whereas no significant association was observed with luminal A subtype and luminal B subtype correlated only to decreased RFS (p=0.0022; Supplementary Figure 2). When the effects of the gene were examined by stage, interestingly ENO-1 overexpression has a good prognostic effect in stage 1 disease and a poor prognostic effect in stage 3 disease (Figure 3). At protein level, there was no correlation of ENO1 expression and survival (p=0.095; Supplementary Figure 3). Although an effect observed at both RNA and protein level is more convincing, many biomarkers that are significant at RNA level are not confirmed to be predictive and/or prognostic at protein level due to post-transcriptional or post-translational modifications affecting protein expression.

**Figure 1.**
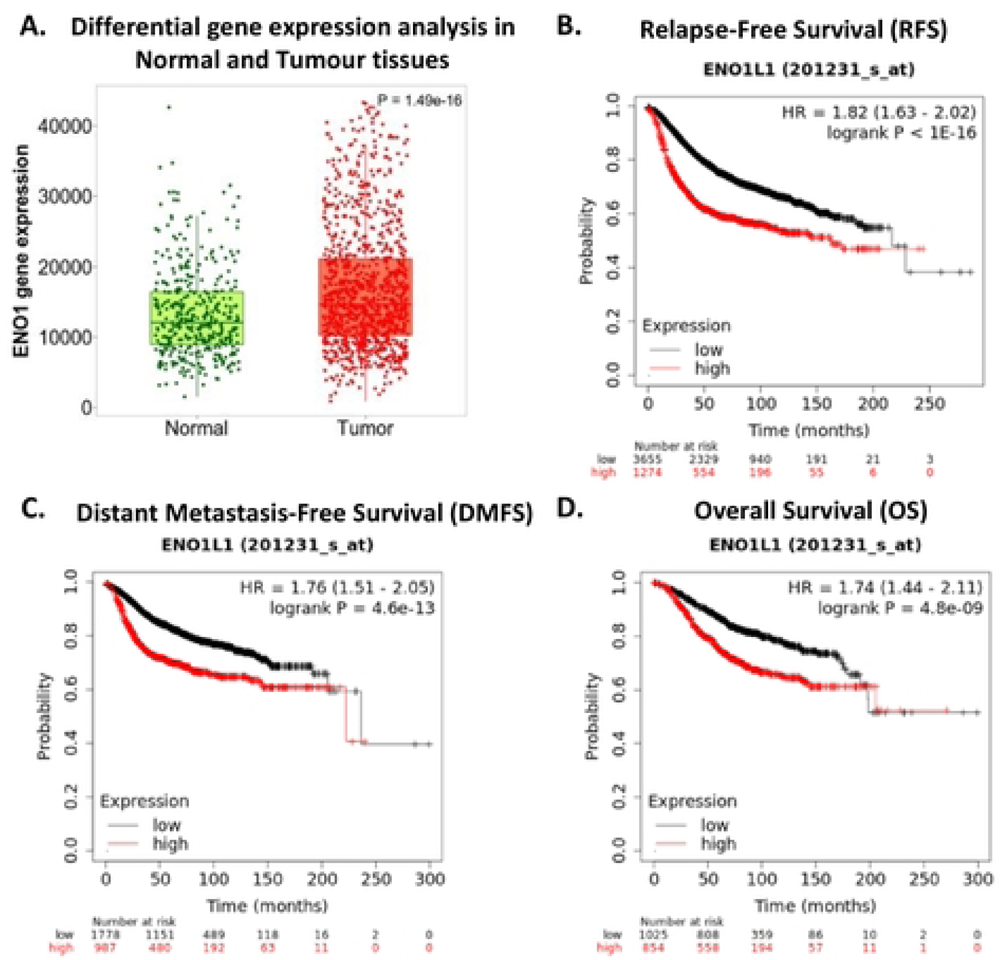

**Figure 2.**
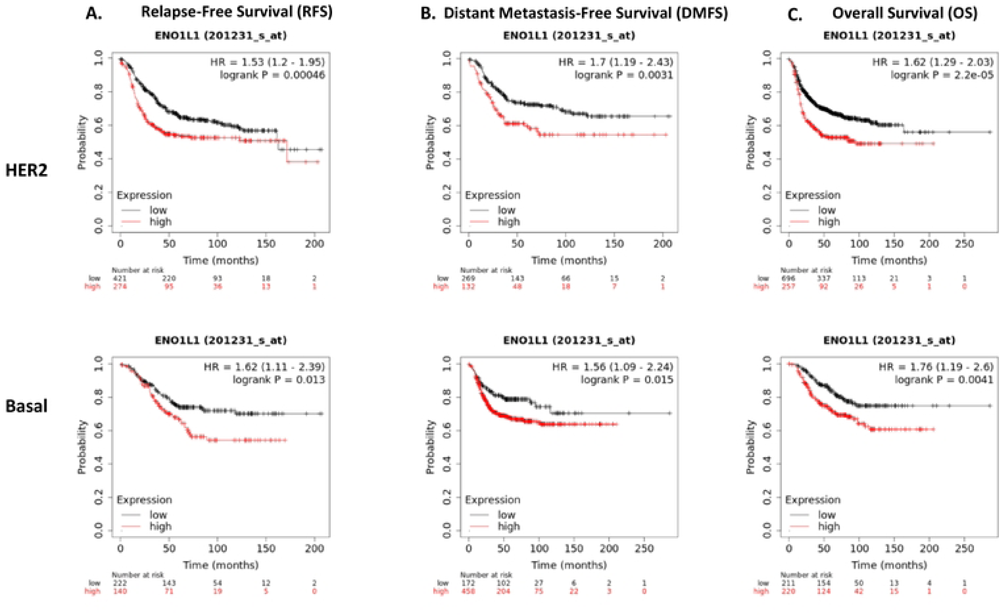

**Figure 3.**
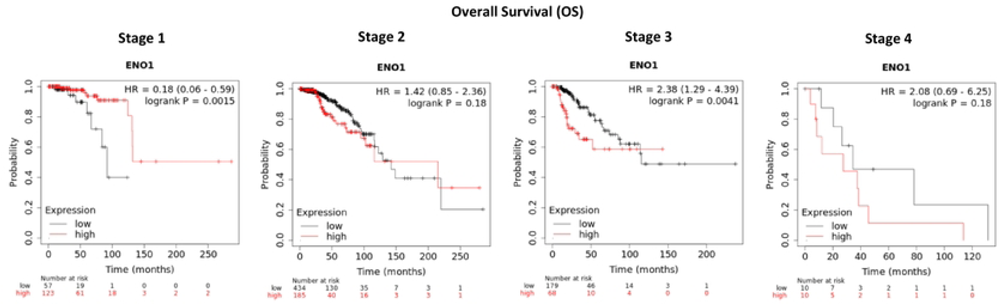

We also carried out an analysis using the ROC plotter tool [29] to investigate whether ENO1 was a biomarker of sensitivity or resistance to endocrine therapy, HER2-directed therapy or chemotherapy in breast cancers. Analysis was performed both in terms of complete pathological response (CPR) and 5-year RFS. There was no correlation between ENO1 and endocrine therapy whereas, a moderate correlation (AUC=0.652, p=0.034) was observed in RFS in patients treated with anti-HER2 therapy (Figure 4A,B respectively). However, in chemotherapy treated patients (Figure 4C), there was moderate correlation with both CPR (AUC=0.541, p=0.032) and 5-year RFS (AUC=0.611, p<0.0001). Our data highlights that patients with high ENO1 expression are or can develop resistance to chemotherapy.

**Figure 4.**
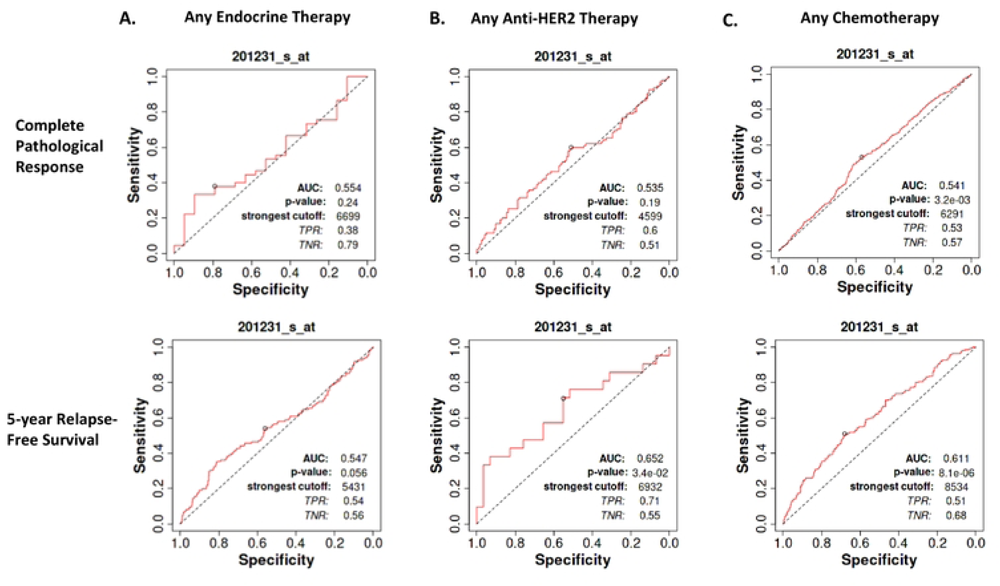

### Genomic alterations in ENO1

Data from cBioportal showed that genomic alterations in the ENO1 gene were detected in 0.2% of breast cancers and they mainly involved copy number alterations (CNA; gene amplifications and deletions). The breast cancer cohorts and the data are presented in Figure 5A. Interestingly, analysis of the metastatic breast cancer cohorts (Figure 5B) showed 7% genomic alterations (gene amplifications, deletions missense and truncating mutations) highlighting that alterations in the ENO1 gene are acquired during treatment and could be related to disease progression and metastasis.

**Figure 5.**
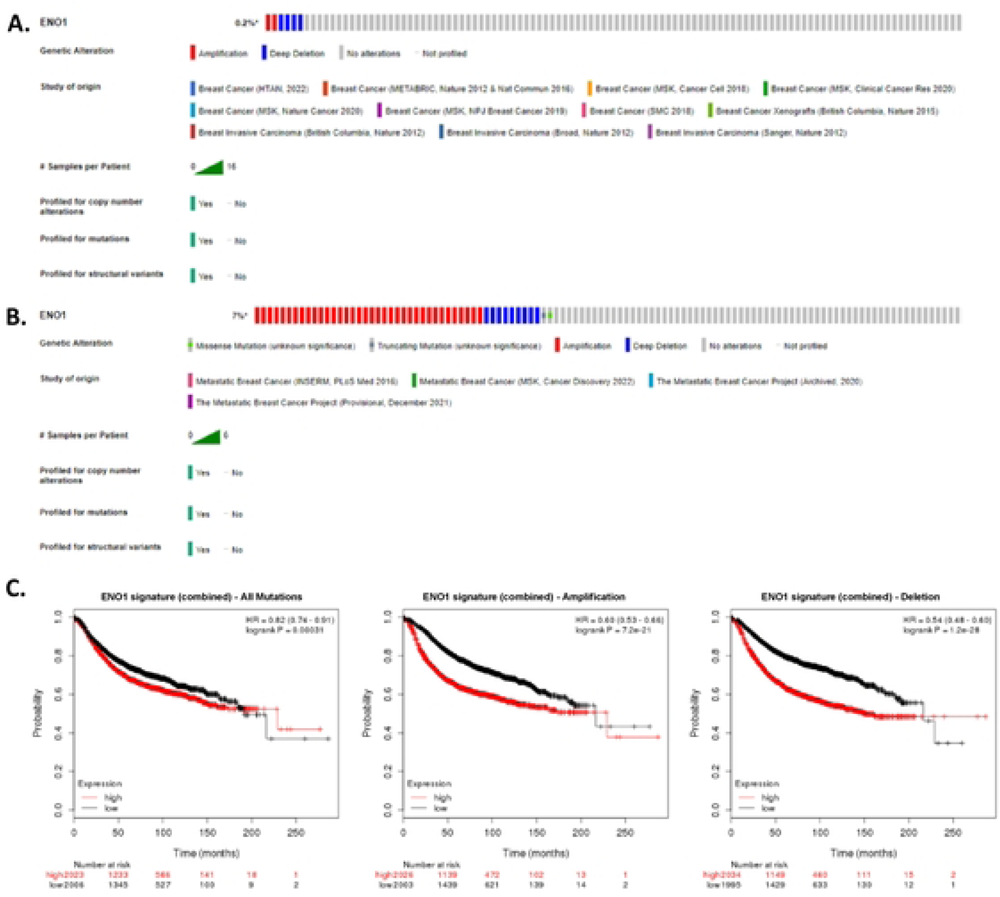

The metagene signatures associated with ENO1 alterations predicted a poor clinical outcome for all types of alterations (mutations, amplification and deletion; Figure 5C).

### Oncomine Network analysis

Nine breast cancer patient cohorts had data available on the Oncomine platform (Supplementary Table 1). Network analysis of the main cancer hallmarks and their associated pathways identified that ENO1 clusters with many signalling, epigenetic and immune pathways. Analysis by receptor status revealed ENO1 overexpression in the triple-negative subtype for all the analysed pathways. In the HER2-positive, overexpression was linked to cytotoxicity, epithelial-to-mesenchymal transition (EMT), histone phosphorylation, costimulatory and cytokine/chemokine signalling, whereas underexpression was linked to hypoxia, MAPK signalling, histone deubiquitination and transcription factor. The hormone receptor-positive subtypes showed ENO1-associated underexpression of all the analysed pathways. All the data is summarised in Figure 6A and Supplementary Table 2.

**Figure 6.**
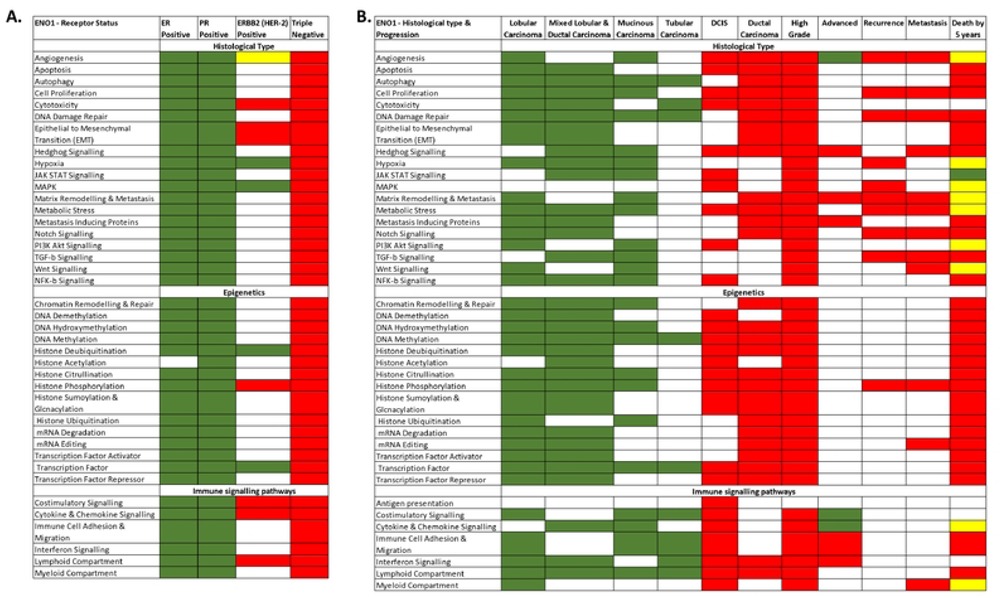

Analysis by histological subtypes and disease progression show that ENO1 was underexpressed in the tumours with better prognosis (Figure 6B). In the lobular and mixed lobular and ductal carcinomas ENO1 was underexpressed for almost all signalling, immune and epigenetic pathways. It was also underexpressed in the rare mucinous and tubular carcinomas is several pathways including autophagy, DNA damage repair, DNA methylation, transcription factor, costimulatory signalling, lymphoid compartment and immune cell adhesion and migration (Figure 6 B).

In the more aggressive ductal carcinoma, ENO1 is overexpressed and clusters with angiogenesis, apoptosis, cell proliferation, cytotoxicity, several signalling and epigenetic pathways in the early stage ductal carcinoma *in situ* (DCIS) whereas more pathways are recruited in ductal carcinoma and high-grade tumours. Interestingly, ENO1 shows clustering with all of the major immune pathways in DCIS and high-grade tumours but only with interferon signalling and lymphoid compartment in ductal carcinoma. There is a general pattern for the clustering to occur in the context of ENO1 gene overexpression as the disease progresses to high grade, advanced, recurrence and metastasis with pathways recruited or disappearing highlighting the importance of ENO1 in the signalling, epigenetic and immune pathways at the different stages. A mixed profile is observed with ENO1 clustering and survival (death by 5 years) where underexpression clusters with JAK-STAT signalling and overexpression with all the epigenetic pathways, immune cell adhesion and migration, Notch, TGF-b, NFK-b, hedgehog signalling and metastasis inducing proteins amongst others. All the data according to histological types and progression is summarised in Figure 6A and Supplementary Table 3.

Methsurv was used to access the effect of ENO1 DNA methylation on survival. Four loci were identified where low methylation is linked to worse survival outcome (Figure 7; cg20971527: HR 0.461, CI 0.312-0.681, LR-test p=0.00014; cg09819654: HR 0.48, CI 0.291-0.792, LR-test p=0.0021; cg06972019: HR 0.662, CI 0.446-0.981, LR-test p=0.038; cg13785123: HR 0.594, CI 0.36-0.978, LR-test p=0.031). All the data is presented in Supplementary Table 4

**Figure 7.**
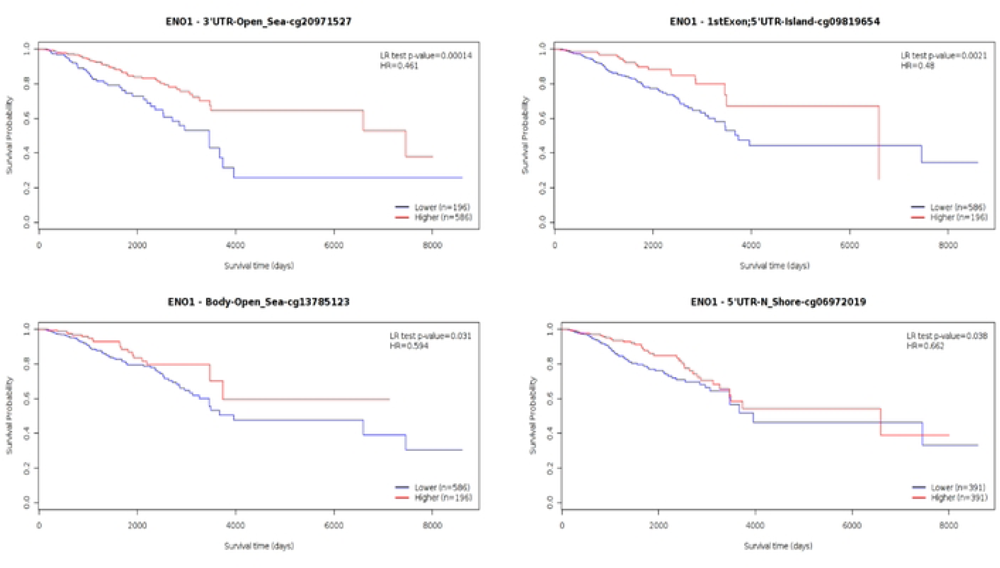

### Immune markers

A low to moderate positive correlation was observed between ENO1 expression and certain tumour infiltrating lymphocytes (TILs), immunomodulators and chemokines whereas a negative correlation was observed with ENO1 methylation. ENO1 CNAs had no effect on the immune markers analysed in TISIDB. In the TILS, the strongest correlations with ENO1 expression were observed with activated dendritic cells (Act-DCs), Gamma Delta T cell (Tγδ), CD56 Natural Killer (NK) cells and activated CD4 T cells-(Act_CD4). Methylation also showed a strong negative correlation with NKT cells, Act_DCs, Tγδ, T helper 1 (Th1) and T helper 2 (Th2) cells, Act_CD4 and activated B cells (Act_B).

A low to moderate positive correlation was identified between expression of ENO1 and expression of immuno-inhibitory genes such as IDO1, IL10RB, LAG3, except PVRL2 that showed a negative correlation to ENO1 expression. Methylation of ENO1 was negatively associated with IDO1, BTLA, CTLA4, IL10, IL10RB, LAG3, TIGIT but positively correlated to PDCD1LG2 and PVRL2. Similar profile was observed with most of the chemokines that negatively correlated with ENO1 methylation and to a lesser extend show a positive correlation to ENO1 expression. The correlation data is analytically presented in supplementary table 5 and examples of the correlation plot of immune markers and ENO1 expression/methylation is presented in supplementary figure 4.

When prognostic data were split by enrichment or depletion of immune compartments, the good prognostic effect of ENO1 in Stage 1 disease was found to be dependent on enrichment of eosinophils, natural killer T-cells, and type-2 T-helper (Th2) cells, and depletion of macrophages. The poor prognostic effect of ENO1 at stage 3 was dependent on enrichement of CD4+ memory T-cells, Natural killer T-cells and eosinophils and depletion of basophils, B-cells, CD 8+ T-cells, macrophages, mesenchymal stem cells and both Type-1 T-helper (Th1) and Th2 cells (Figure 8). Similarly, enrichment of basophils, B-cells, CD8+ T-cells, regulatory T-cells and depletion of eosinophils, CD4+ memory T-cells and Th1/Th2 in association with high mutation burden conferred poor prognosis (Figure 8).

**Figure 8.**
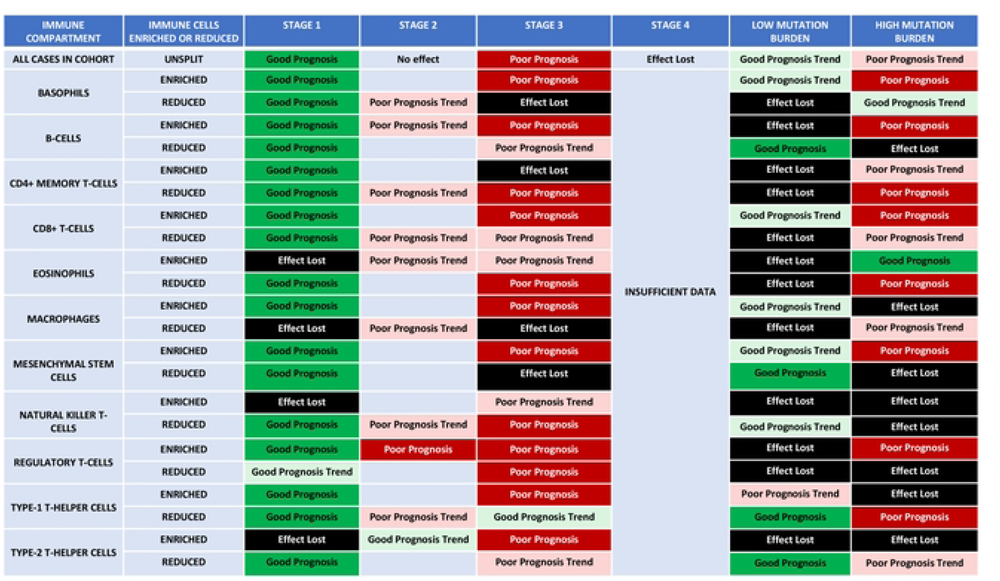
Immune System Interaction with ENOl.

## Discussion

This study has revealed that ENO1 is a poor prognostic marker at RNA level, particularly in triple negative breast cancer. The poor prognostic effect is observed in advanced (Stage 3) disease, and is present in HER-2 positive and triple negative breast cancers. Our data agrees with previous recent studies showing an association of high ENO1 expression with the aggressive basal subtype and a favourable prognosis in patients with early stage breast cancer but not with advanced stage disease and/or basal breast cancer [16,26,38]. We also observed that methylation of the gene is associated with a good prognostic effect, consistent with the expected effect from reducing ENO1 gene expression. In addition to methylation, the gene clusters with a wide range of epigenetic pathways, and this effect is established at the *in-situ* stage.

Although genomic alterations of ENO1 in breast cancer are rare, an increased rate of ENO1 alterations in metastatic tumours as compared with primaries was identified, indicating that ENO1 alterations accumulate with progression and treatment. Both amplifications and deletions were detected, and both are associated with a poor prognosis, providing a partial explanation for the emergence of the poor prognostic effect in more advanced disease. ENO1 clusters with overexpression of all of the pathways involved in cancer hallmarks and epigenetic regulation in triple negative breast carcinomas. We observed a tendency to recruit these pathways in the context of high-grade disease, ductal carcinoma and tumours with a poor 5-year survival. By contrast, ENO1 is a good prognostic marker in stage 1 disease, and clusters in the context of underexpression with a wide range of the pathways in subtypes of breast carcinoma known to be associated with a good prognosis, such as mucinous and tubular carcinomas. Clustering of ENO1 across the pathways we examined was also observed in DCIS, indicating that the gene plays a role from the earliest stages of breast cancer development.

A complex relationship of ENO1 with immune pathways was revealed by this study to complement other studies evaluating the role of ENO1 in the tumour-immune microenvironment [13,38,39]. Underexpression occurs in good prognostic subtypes of breast carcinoma, whereas overexpression of the pathways occurs in triple negative breast carcinomas. Overexpression of the gene tended to promote immune inhibitors and suppress chemokines. However, despite this, higher expression of the gene correlated with increased infiltration of TILs, whereas methylation of the gene was associated with decreased infiltration of TILs, likely reflecting the dominating influence of the broader immune pathway recruitment observed in the network analysis. There will likely be functional effects of ENO1 overexpression, such as a reduced pH of the microenvironment caused by increased glycolysis, which would also tend to promote immune cell recruitment. Splitting survival data by stage and immune compartment revealed complex effects, with the poor prognostic effect at stage 3 being dependent on enrichment or depletion of a wider range of immune cells than the good prognostic effect seen in stage 1. The stage 1 good prognostic effect tended to be dependent on depletion of a narrow range of immune pathways, whereas the poor prognostic effect at stage 3 was dependent on enrichment of a wider range of different immune pathways, likely reflecting the ability of the tumour to co-opt the immune system. These data overall point to a potential role of ENO1 in recruiting immune pathways, particularly in triple negative breast carcinomas and advanced disease, immediately suggesting that there may be a role for ENO1 targeted therapy as an adjunct to immune therapy.

## Conclusions

This large scope analysis highlights further the potential of ENO1 as a novel biomarker of breast cancer progression and in particular in the triple-negative/basal subtype, a subtype with worse prognosis than the hormone-positive and the HER2-positive subtypes. Overall, ENO1 is overexpressed in comparison to normal tissue and confers a worse prognosis in breast cancer. Broad epigenetic associations are established by the time DCIS has become invasive and more are observed as the cancer progresses up to death. In addition, more pathways involved in the cancer hallmarks and the immune pathways are recruited. The observations in this study warrant further investigation to characterise better the relationship of ENO1 with epigenetic and immune-related pathways. This will help the development of therapeutics targeting ENO1 directly and/or as an adjunctive treatment to immunotherapies.

## Data Availability

All data generated or analysed during this study are included in this article and its supplementary information files. The datasets supporting the conclusions of this article have been generated using the following websites: www.kmplotter.com, http://www.g-2-o.com http://www.cbioportal.org https://www.oncomine.org https://biit.cs.ut.ee/methsurv/ www.rocplotter.com https://epifactors.autosome.ru http://cis.hku.hk/TISIDB/ and appropriate references to these are included in the manuscript.

## Acknowledgements

The authors would like to thank the bioinformatics department at the University of Liverpool who have approved the processes to produce this manuscript.

## Ethics approval and consent to participate

Not applicable

## Consent for publication

Not applicable

## Availability of data and materials

All data generated or analysed during this study are included in this article and its supplementary information files. The datasets supporting the conclusions of this article have been generated using the following websites: www.kmplotter.com, http://www.g-2-o.com; http://www.cbioportal.org; https://www.oncomine.org; https://biit.cs.ut.ee/methsurv/; www.rocplotter.com; https://epifactors.autosome.ru; http://cis.hku.hk/TISIDB/ and appropriate references to these are included in the manuscript.

## Authors’ contributions

Data obtained by Dr Vijay Sharma, Dr Alistair Heath and Dr Athina Giannoudis. AG and VS have analysed the data and produced the body of the manuscript. All authors have read and approved the submitted manuscript.

